# Is anybody ‘Learning’ from Deaths? Sequential content and reflexive thematic analysis of national statutory reporting within the NHS in England 2017-2020

**DOI:** 10.1101/2022.07.12.22277526

**Authors:** Z Brummell, D Braun, Z Hussein, SR Moonesinghe, C Vindrola-Padros

**Affiliations:** University College London; Advisor/Lived experience

## Abstract

**Introduction:** The imperative to learn when a patient dies due to problems in care is absolute. In 2017, the Learning from Deaths (LfDs) framework, a countrywide patient safety programme, was launched in the National Health Service (NHS) in England. NHS Secondary Care Trusts (NSCTs) are legally required to publish quantitative and qualitative information relating to deaths due to problems in care within their organisation, including any learning derived from these deaths.

**Method:** All LfDs report from 2017 – 2020 were reviewed and evaluated, quantitatively and qualitatively using sequential content and reflexive thematic analysis, through a critical realist lens.

**Results:** The majority of NSCTs have identified learning, actions and, to a lesser degree, assessed the impact of these actions. The most frequent learning relates to missed/delayed/uncoordinated care and communication/cultural issues. System issues and lack of resources feature infrequently. There is significant variation amongst NSCTs as to what ‘learning’ in this context actually means and a lack of oversight combining patient safety initiatives.

**Discussion:** Engagement of NSCTs with the LfDs programme varies significantly. Learning as a result of the LfDs programme is occurring. The significance or value of this learning in preventing future patient deaths remains unclear. Consensus about what constitutes effective learning with regards to patient safety needs to be defined and agreed upon.

## Introduction

Within higher income countries, between 0.5% and 8.4% of hospital deaths are preventable or potentially preventable.[1] The need to learn, to disseminate learning and to demonstrate that learning has been achieved when a patient dies due to problems in care is highlighted in multiple patient safety reports and inquiries both nationally and internationally.[2-5] Bereaved families frequently voice that learning to prevent future harm, in addition to acknowledgement and support for their harm are needed.[6-8]

In 2017, the Learning from Deaths (LfDs) framework, a countrywide patient safety programme, was launched in the National Health Service (NHS) in England. This was in response to reviews at Southern Health NHS Trust and then other NHS Secondary Care Trusts (NSCTs) within England demonstrating a lack of systematic approach and meaningful change occurring in response to unexpected deaths.[9-11] Guidance was subsequently published on implementing the LfDs framework at NSCT board level,[12] with amendments to statutory regulations, making annual reporting of both quantitative and qualitative information relating to patient deaths a legal requirement in England.[13] The quantitative information required included the number of patients who died more likely than not due to problems in care. The qualitative information required included a description of learning, actions taken as a result of learning and an assessment of impact of these actions in relation to deaths due to problems in care. The reporting mechanism was built into the NHS ‘Quality Accounts’ system: where NSCTs are legally required to produce a publicly available annual report about the quality of their services (UK government legislation).[14] Guidance was given that NSCT board leadership should ‘share relevant learning across the organisation and with other services’,[12] and that NSCTs should ‘engage meaningfully with bereaved families and carers’.[8] There was no guidance about the format in which the qualitative data should be presented or suggested methods NSCTs could use to evaluate impact. Lalani and Hogan (2021), in their narrative account of the key drivers in the development of the LfDs programme, highlighted the tension arising due to competing goals of developing a learning culture while also increasing accountability through regulation.[15]

Prior to the introduction of the LfDs framework, there has been a longer standing legal requirement within the United Kingdom (UK), that all deaths including NSCT deaths where the cause is unknown, or of unnatural cause are referred to and investigated by a Coroner. Coroners are statutorily required to issue a Prevention of Future Death (PFD) report to individuals or organisations who are able to action appropriate changes where problems in care are identified as potentially causing or contributing to a death.[16] Leary et al (2021) review of PFD reports from 2016 – 2019, found five common learning ‘themes’: A deficit in skill or knowledge, missed/delayed/uncoordinated care, communication/cultural issues, system issues or lack of resources.[17] The ‘themes’ identified (inductively) are similar to those identified in many other UK inquiries and reports from organisations investigating deaths due to problems in care.[18-20]

Historically, recording, investigating, reviewing incidents and superficial learning appear to have been the major focus in patient safety, with less focus on effective learning through action, demonstrating the real impact of this intervention and spreading adoption when impact is found.[21] For example, with PFD reports, NSCTs are expected to respond (within 56 days) explaining how they intend to or have made changes, however there is no process for following up actions proposed, including decisions to take no action, or for assessing whether any actions taken have resulted in measurable improvements.[16] Another example is found in the 2019 CQC report which reviews the ‘first year’ of NSCTs implementing the LfDs guidance, where four of the five future recommendations in the report focus on the process of investigation.[22] Investigating while important is not in itself going to enable discovery of effective solutions, NSCTs and NHS staff often know what the problems are, but don’t have the resources or impetus to fix them.

This study thematically analyses LfDs reports to understand what we can learn from reported ‘learning’, what actions NSCTs are taking in response to this learning, how NSCTs are assessing the impact of these actions and overall engagement with the LfDs programme. In addition, we analyse two non-statutory requirements of the LfDs reports: sharing of learning and engagement with bereaved families and carers. This study does not analyse the quality of reporting, this will be presented in a separate paper.

## Method

This is a study of an NHS safety improvement programme positioned through a critical realist lens to identify and understand the real causal mechanisms enabling or preventing engagement with the LfDs programme.[23] We undertook a secondary source document analysis using sequential content (for quantitative counts through a qualitative process), then thematic (for fuller qualitative) analysis of all LfDs reports from NSCTs between 2017 – 2020 (ambulance Trusts were excluded as they weren’t required to report in 2017/18).[24, 25] Content analysis as described by Morgan (1993) is used to describe and quantify “patterns in the data”.[26] Reflexive Thematic Analysis (TA) as described by Braun and Clarke (2021) has been used for its flexible, yet structured approach to find “shared meaning” in the data while enabling researcher consideration of their own impact on data interpretation.[27] Reflexive TA in this study enables evolution of and contextualisation of the content analysis through greater understanding of the causal mechanisms affecting engagement with the LfDs programme. The authors have purposefully used these two different approaches, despite the tensions arising, to provide a fuller explanation of NSCTs empirical reporting, to engage with frontline staff (who embrace quantitative research) and to reveal underlying deeper causal mechanisms impacting engagement with health policy. Our objectives for this study were to derive key understanding for the NHS frontline staff, managers, policymakers and beyond, by analysing LfDs reports.

A deductive approach to content analysis was undertaken, informed by Brummell et al (2020) paper and previous research and enquiries analysing deaths due to problems in care as described in the introduction.[1] Content analysis, then reflexive TA, were sequentially used to analyse: learning, actions taken as a result of learning, an assessment of impact of these actions, how bereaved families had been involved in any learning and if/how learning had been shared. With reflexive TA a deductive then subsequent inductive approach was used to assess NSCT engagement with the LfDs programme. The methodology used was a process of data familiarisation (reading each report twice on separate occasions), systematic data coding (through content analysis), generating initial domains deductively, then developing multi-dimensional themes inductively through active engagement and immersion in the data, looking both at what was present and what was absent. Finally, these themes were refined by sense checking with the original reports and discussion with the other authors.

Every LfDs report was reviewed by the primary reviewer (ZB) twice, on separate occasions to ensure full data capture. 10% of reports from 2018/19 and 2019/20 were identified by random number generation and reviewed independently by a second reviewer (ZH) to ensure accurate and reliable coding for the content analysis. In the case of disagreement, (of which there was only one for each year) ZB re-reviewed the LfDs report for clarification. Reflexive TA was undertaken by ZB only.

Data were captured in Microsoft excel (V.16.15). This study has been reported using Standards for Reporting Qualitative Research, for the content analysis.[28]

### Reflexivity

Reflexivity was undertaken alongside the research methodology, as described by Trainor & Bundon, 2021).[29] For additional information see supplemental page 1.

### Patient and public involvement

This study forms part of a larger programme of work overseen by a public and relatives steering group to improve relevance from the perspective of those affected by deaths in healthcare and to reduce biases from the healthcare staff researchers. The steering group have been involved in the planning, design and development of conclusions, through videoconferencing and email correspondence. The involvement of a steering group member in authoring this paper has significantly and positively influenced the reporting of this study, ensuring focus on reporting family involvement. The reporting of PPI has been undertaken using guidance for reporting involvement of patients and the public 2—short form.[30]

## Results

### Quantitative Content Analysis

The number of NSCTs is reducing year on year due to mergers of NSCTs: 222 NSCTs in 2017/18, 217 NSCTs in 2018/19, 213 NSCTs in 2019/20

Through systematic data coding, different types of ‘learning’ and ‘action’ were identified, defined and refined. The most common ‘learning’ and actions undertaken across all NSCTs (as % of all trusts who reported learning: 2017/18 N = 195, 2018/19 N = 191, 2019/20 N = 173) can be found in Table 1 and Table 2.

**Table 1.** The most common learning found across all NSCTs.

| **Learning** | **NSCTs citing 2017/18 (%)** | **NSCTs citing 2018/19 (%)** | **NSCTs citing 2019/20 (%)** |
| --- | --- | --- | --- |
| **Problem with communication (including language barrier, consent & handover)** | 46% | 45% | 41% |
| **Problem in recognition & escalation of deteriorating patients** | 43% | 46% | 36% |
| **End-of-life planning or treatment escalation planning not evident/incomplete** | 42% | 47% | 45% |
| **Problems with documentation** | 41% | 49% | 47% |
| **Lack of clinical knowledge, consideration differential/delay diagnosis or seeking advise** | 27% | 23% | 27% |
| **Lack of/problem with risk assessments/interventions/escalating concerns** | 25% | 35% | 32% |
| **Lack of/problem with engagement with/support of families/carers** | 4% | 15% | 28% |

**Table 2.** The most common actions undertaken across all NSCTs.

| **Action Undertaken** | **NSCTs citing 2017/18 (%)** | **NSCTs citing 2018/19 (%)** | **NSCTs citing 2019/20 (%)** |
| --- | --- | --- | --- |
| **Review and improve process (including governance, risk, pathway, strategy)** | 67% | 56% | 57% |
| **Highlight or produce or review guidelines/protocols/policies/treatment bundle/toolkits** | 51% | 52% | 46% |
| **Implementation programme of training/ education** | 51% | 44% | 39% |
| **Quality improvement work, audit or similar** | 47% | 50% | 39% |
| **Work to improve communication/collaboration/shared learning** | 43% | 49% | 57% |
| **Improved documentation/coding** | 25% | 39% | 33% |

‘Learning’ and actions undertaken as cited in the LfDs reports were assigned into domains as seen in Figure 1 and Figure 2. Domains are also referred to as ‘topics’ or even as ‘themes’ by some researchers. [31]

**Figure 1.**
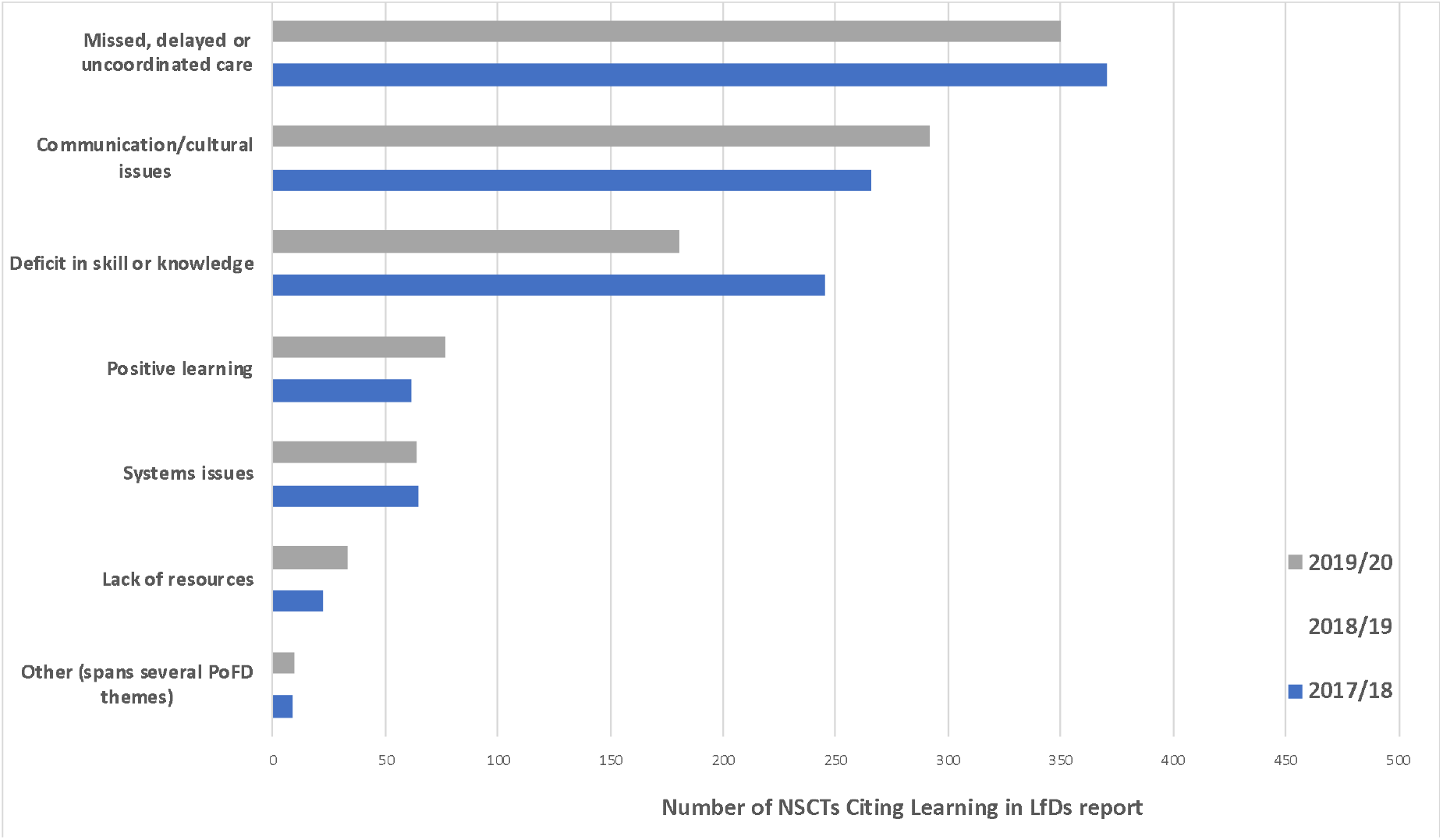
Frequency table of learning domains identified from all NSCTs.

**Figure 2.**
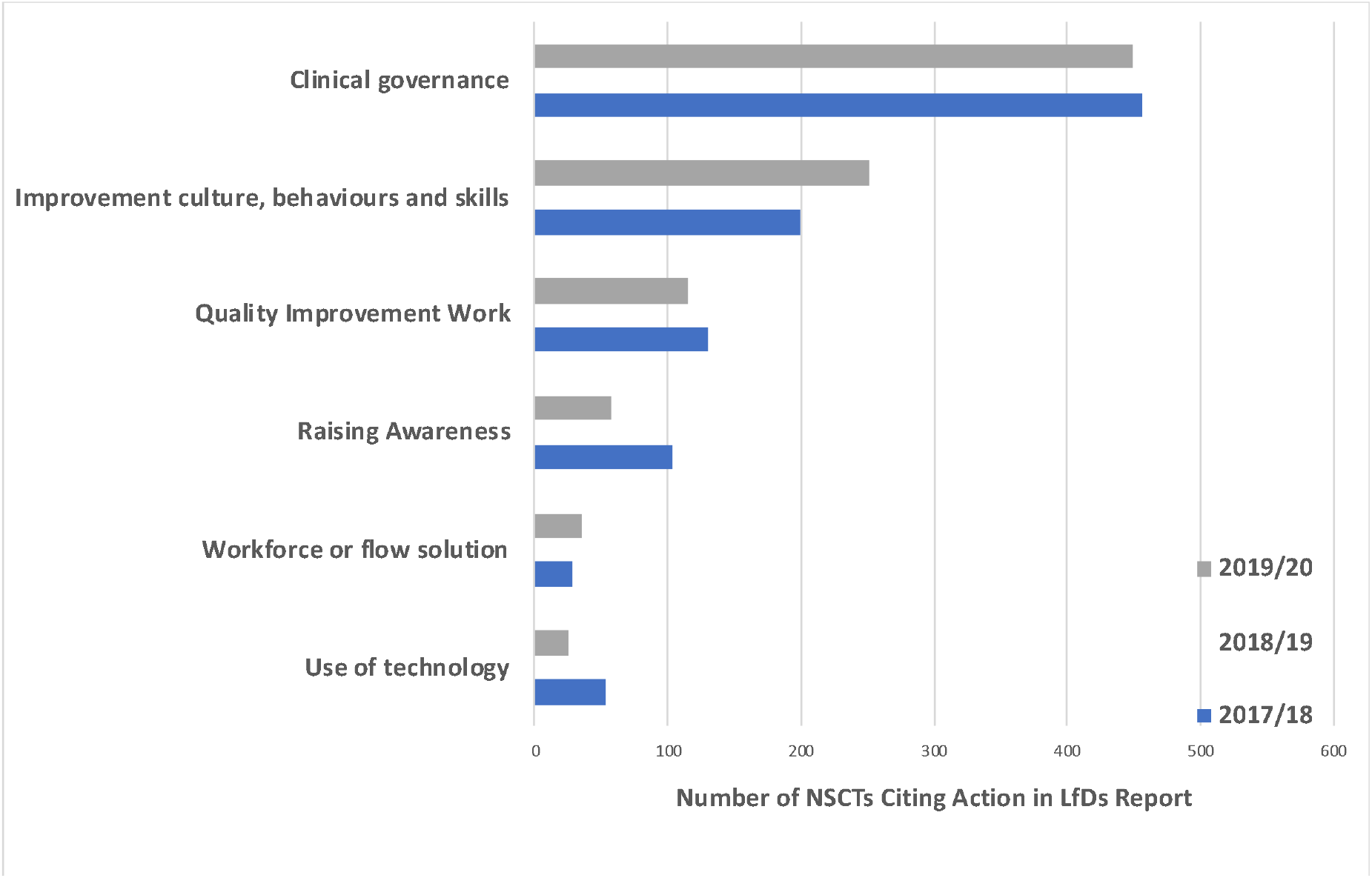
Frequency table of action domains identified from all NSCTs.

### Qualitative Content Analysis

Very few NSCTs articulate the problem or problems in care that led to the reported ‘learning’ and/or actions. Where the problem or problems were identified, there was a lack of detail about the problem to understand what actually happened to the patient and why. Many NSCTs did not differentiate ‘learning’ from actions or confused ‘learning’ with actions. Some NSCTs described ‘learning’ not followed by actions addressing the ‘learning’. NSCTs did not comment on LfDs ‘learning’ or action from previous years and did not describe the impact of this or how things had changed or improved year on year. Many NSCTs who did not report deaths due to problems in care still reported ‘learning’ and/or actions.

#### Assessment of impact of actions

NSCTs who published a LfDs report and provided information about an assessment of impact of actions is as follows:

- In 2017/18: 105 out of 220 (47%)
- In 2018/19: 123 out of 214 (57%)
- In 2019/20: 79 out of 202 (39%)

Assessment of impact of action includes NSCTs that have already assessed impact or have a plan to assess impact, but not NSCTs that state they have assessed impact, but have instead provided unrelated information which does not assess impact.

Some NSCTs have made efforts to thoroughly assess impact with one even differentiating qualitative and quantitative assessment. Many of NSCTs that did assess impact used audits and/or quality improvement projects to do this:

- *‘The organisation has a variety of audit programmes running which will check if any learning from deaths is put into practice’*
- *Use of audit data (for example improvement in TEP & DNACPR completed from 23% to 78% following introduction of a combined form’*
- *‘Within the Forensic Services, the Performance Department provides an ‘action plan/QIP summary & updated action plan/QIP monitoring sheet’ monthly report that the group monitors and reviews’*
- *‘The QI plans that link directly to Sepsis and the Deteriorating patient have just begun. However, the progress of these plans is being monitored in the structure of the measuring and monitoring framework for safety*.*’*

The other major way that impact was assessed was through observing changes in Standardised Hospital Mortality Index and/or Hospital Standardised Mortality Ratio:

- *‘The impact of the case record review process and the associated improvement actions can be assessed using the hospital standardised mortality ratio (HSMR)’*
- *‘This work has now been completed and the Trust has seen an improvement in both its key headline mortality indicators SHMI and HSMR*.*’*

Some NSCTs did not fully specify how impact had been assessed or quantified, but did note that impact has occurred:

- *‘The impact of the project has been notable, and we will now update our policy and procedure to reflect our learning’*
- *‘All actions are monitored to ensure they have had the desired impact. If this has not happened, actions will be reviewed and altered as necessary to ensure that sustainable and appropriate change has been implemented’*
- *‘The Trust believes that by implementing the above actions, patient safety and quality of care has improved’*
- *‘No further incidents have been reported similar to the cases identified above and it therefore appears that these were isolated cases and/or actions taken have been effective*.*’*

Several NSCTs appear to have misunderstood what is meant by assessment of impact, for example reiterating the purpose or process of the LfDs programme:

- *‘Perhaps the most important impact continues to be raising of awareness, amongst Trust staff, of the importance of reporting deaths through our incident reporting system as a potential learning opportunity’*
- *‘A key impact is the need to develop our work with mortality reviews during 2018/19 so that all reviews are consistently undertaken to a high standard’*
- *‘The impact has been to continue to share and embed the learning identified from ongoing review of deaths, including people with learning disabilities and therefore improve patient care*.*’*

Or issued statements that don’t entirely make sense:

- *‘Where the greatest impact is evidenced is through the analysis of themes and trends so amalgamating the learning from mortality reviews with outcomes from other investigations such as complaints, serious incidents, Never Events and legal claims to identify the full range of issues is proving essential’*
- *‘It is anticipated that the impact of the actions and learning described above will impact on the care provided to patients in receipt of services across the Trust*.*’*

Or made statements that do not assess impact of their actions despite report headings suggesting they are:

- *‘The Trust is 98% compliant with all relevant recommendations outlined within NICE guideline for sepsis, with the exception of three recommendations which relate to the provision of information leaflets for adults, this work is currently in progress*.*’*

Several NSCTs gave reasons for not including an assessment of impact:

- *‘An assessment of the impact of the actions taken as a result of the completed reviews will be included in each report to the Board, as the completed reviews are still under review by the services the impact cannot be described at this stage’*
- *‘It is difficult to determine the impact of lessons from multifactorial events such as these where human factors around decision making and communication are the root cause’*
- *‘It is not possible to comment on the effect of most these actions in the time period under consideration’*
- *‘The actions referred to in 27*.*5 have either only recently been completed or remain outstanding and due for completion in 2020/21, therefore it is not possible to make an assessment of the impact of the action*.*’*

Or describe the difficulties in assessing impact of actions:

- *‘Due to the scale of work involved in the areas listed it would be premature to evaluate the outcomes’*
- *‘This is difficult to quantify as the learning has been gradually developed and shared throughout the year’*
- *‘It is very difficult to assess the impact resulting directly from mortality reviews due to a number of different factors. These include:*
  - *challenges with co-ordinating local (directorate) governance and learning with Trust-wide processes*.*’*

#### Involvement of bereaved families

Of the NSCTs who published a LfDs report, those that mentioned the involvement of families/carers either in the investigation process or in shared learning or that they communicate with/support/engages families/carers after a patient dies:

- In 2017/18: 37 out of 220 trusts (17%)
- In 2018/19: 80 out of 214 trusts (37%)
- In 2019/20: 90 out of 202 trusts (45%)

Several NSCTs demonstrated significant positive engagement with families, particularly in 2019/2020, often (but not always) through the Medical Examiner system:

- *‘Promote and support involvement of patients’ families in investigations’*
- *‘Improve mortality reviews and embed the new medical examiner process, providing families, carers and staff with opportunities to both raise concerns and highlight examples of good practice and excellent care’*
- *‘Changes have been made to investigation templates and staff have been trained in ‘Patient Liaison’ by the Health Safety Investigation Branch. Families concerns are now identified and included in Stage 3 mortality reviews’*
- *‘The Trust continues to ensure that our direct contact with any family affected by the death of a loved one is paramount in our processes of learning from deaths, giving each family the opportunity to contribute to the Terms of Reference for an investigation, comment on a draft report and have full disclosure of the report at sign off by the Trust*.*’*

It is clear that by 2019/20 several NSCTs have significantly developed their Medical Examiner system. Many of these NSCTs describe in detail the ME process, but omit other statutory requirements. Another way in which NSCTs are supporting bereaved families is through specific roles, such as Family Liaison Officers, bereavement midwives and Bereavement Liaison Practitioners: ‘*In December 2018 the Trust introduced a Bereavement Liaison Practitioner. Within the first 12 month of the Bereavement Practitioner commencing in post approximately 250 individual staff members or family members were supported’*

#### Sharing Learning

Of the NSCTs who published a LfDs report, those that have shared or plan to share the learning more widely within their own organization:

- In 2017/18: 106 out of 220 (48%)
- In 2018/19: 90 out of 214 (42%)
- In 2019/20: 93 out of 202 (46%)

This sharing occurs through a variety of communication mediums: Face to face meetings, emails, screensavers, newsletters, events, intranet (case studies, safety alerts), for example:

- *‘The Trust has led two health economy-wide Learning from Death events to ensure learning and improvement is shared across organisational boundaries’*
- *‘Sharing of the learning from mortality reviews continues both internally and externally, with Trust staff presenting to local GP training sessions’*.

Of the NSCTs who published a LfDs report, those that have shared or plan to share the learning outside their organisation, with neighbouring NSCTs, Clinical Commissioning Groups or other national organisations:

- In 2017/18: 17 out of 220 (8%)
- In 2018/19: 28 out of 214 (13%)
- In 2019/20: 20 out of 202 (10%)

### Reflexive Thematic Analysis

Through reflexive thematic analysis the following themes were identified:

- *‘What does ‘learning’ mean?’*
- *‘Undertaking a thematic analysis’*
- *‘Opportunities to triangulate’*
- *‘Feeling the pressure’*
- *‘Description of the incident/problem’*
- *‘The importance of culture’*

Further detail can be found in supplemental page 2.

## Discussion

There is no consensus as to what ‘Learning’ in the context of patient safety actually means. ‘Learning’ or to ‘Learn lessons’ appears as a catch-all term which NSCTs recognise they must do or be seen to be doing following a patient safety incident and is a frequent NSCT quote in broadcast media.[33-34] This lack of definition, understanding and consensus of what is meant by effective learning by healthcare organisations may be one of the main barriers to sustained improvement in patient safety. Knowledge sharing, and effective learning are complex social activities,[35] made even more complicated when undertaken in complex organisations with transient workforces, reduced resources and competing priorities.[36] A greater understanding of core concepts in ‘Learning’ such as double-loop learning, and the factors required for successful organisational learning need to be considered when developing healthcare safety policy.[15, 37]

An important issue to highlight with regards to ‘Learning’ is the discord between NSCT intention of ‘learning’ and a lack of effective actions, potentially resulting in public distrust of healthcare professionals and organisations, with the propagation of harm to bereaved relatives. This lack of ‘learning’ is noted by Leary et al (2021) where they comment that Coroners ‘*expressed concern or even frustration that learning or actions from PFDs was not utilised and that organisations repeatedly appeared before them*.’[17]

In this study and Leary et al (2021), systems/lack of resources are relatively infrequently cited as being the problems relating to deaths due to problems in care.[17] This is at odds with the patient safety literature and of that of healthcare investigation organisations.[38-39] Linked to this is the lack of recommendations from NSCTs for improvements that focus on systemic change and redesign rather than individual performance. From the LfDs reports, it is not clear if NSCTs are not able to recognise system failure (which seems more likely) or if systems/lack of resources are not an issue. What is also missing from LfDs reports is the triangulation of data and recognition of overlapping patient safety programmes such as ensuring a ‘just culture’.[40] Leadership teams in NSCTs and organisations regulating healthcare should recognise and understand patient safety as a whole, not as a series of discrete policies/programmes. Integrating instead of layering should improve both the success of these policies/programmes and the overall functioning of NSCTs.

Missed/delayed/uncoordinated care and communication/cultural issues are the two major domains cited in ‘learning’. These two domains were also in the top three ‘themes’ identified by Leary et al (2021) and align with the three recurring patient safety ‘themes’ identified by the Healthcare Safety Investigation Branch in their thematic review (2021). [17, 39] In this study, we found ‘learning’ codes were sometimes overlapping domains, for example demonstrating both deficits in the skill/knowledge in combination with missed/delayed/uncoordinated care. With regards to the specific ‘learning’ codes derived from this study, there is a reduction in reporting of ‘Problem in recognition & escalation of deteriorating patients’ from 46% in 2018/19 to 36% in 2019/20. This may signify an improvement and/or effective learning and/or actions occurring related to this area of practice. This would fit with the increased UK national focus around recognition of the deteriorating patient. [40, 41] Perhaps a similar approach could be undertaken for enabling improvement with other frequently raised ‘learning’ codes such as ‘problems with documentation’; through an increased national focus around documentation, clinical coding and integrated electronic health records.[42, 43]

There was an increase in reporting of ‘Lack of/problem with engagement with/support of families/carers’ from 4% in 2017/18 to 28% in 2019/20. This could be due in part to NSCTs increasing awareness of the LfDs guidance for NHS trusts on working with bereaved families and carers,[8] more likely though is because of the increased establishment of Medical Examiner Systems supporting the bereaved within NSCTs.[44] Further recognition of the need for and provision of training for healthcare staff with regards to bereaved relatives was identified by a few NSCTs. Despite evidence that patient and family engagement improve patient safety, many organisations still have difficulties achieving this.[45]

In the case of action, this study demonstrates a reduction in reporting of ‘Review and improve process (including governance, risk, pathway, strategy)’ from 67% in 2017/18 to 57% in 2019/20 and a reduction in reporting of ‘Implementation programme of training/ education’ from 51% in 2017/18 to 39% in 2019/20. The reason for these reductions could be that these actions were implemented in 2017/18 and therefore NSCTs have not reiterated these actions in subsequent years. There is some evidence that teaching and training initiatives such as clinical simulation are credible improvement methods.[46] There was increased reporting of ‘Work to improve communication/collaboration/shared learning’ from 43% in 2017/18 to 57% in 2019/20. This may reflect the increasing understanding in the critical importance of knowledge transfer and knowledge management for patient safety.[47, 48] Approximately half of all NSCTs did demonstrate shared learning within NSCTs, through a variety of platforms, there was however very limited evidence of shared learning between organisations. Karam et al (2018) found interorganisational collaboration in healthcare to be more challenging than interprofessional collaboration.[49] This lack of interorganisational collaboration could reflect the enduring effect of the internal market, despite subsequent changes to amend this.[50, 51] A lack of system-level thinking as described by Vindrola-Padros et al (2020) is evident throughout the LfDs report, in actions implemented and ‘learning’ shared.[52] Clinical governance is the major domain cited as an action in the LfDs reports. The ability of NSCTs to undertake effective governance is variable, affected by multiple factors including communication and relationships between Clinicians and Managers.[53, 54]

Concerningly assessment of impact of actions reduced significantly in 2019/20, after an initial rise between 2017/18 and 2018/19. Whether this effect was related to resourcing issues for data collection and reporting relating to COVID-19 is unknown. This reduction in assessment of impact could signify a loss of impetus for the LfDs programme. NSCTs appear to need further assistance in meaningfully assessing impact and that the ability to do so would be of global benefit to NSCTs, out with the LfDs programme. Specific guidance for patient safety teams within NSCTs in how to evaluate what are often complex interventions, could be adapted from the UK Medical Research Council and National Institute of Health Research framework for the development and analysis of complex interventions.[55]

There is ongoing variable NSCT engagement with the LfDs programme. Inconsistencies in LfDs and PFD reporting raise questions about what real world difference statutory reporting makes. Understanding the real causal mechanisms/factors enabling or preventing engagement with the LfDs programme needs to be understood for success of this and future health policies and legislation. The consequences of not LfDs are multiple: economic, [56] reputational, human and moral.

The aspiration of organisational memory does not seem possible in many NSCTs currently. [57, 58] However some NSCTs appear to be exceling in patient safety despite similar constraints, having developed a strong patient safety culture through continuous improvement.[59] The development of a culture of safety is central to any sustainable efforts towards patient safety improvement.

## Conclusions

Engagement of NSCTs with the LfDs programme varies significantly. Some NSCTs are undertaking ‘learning’ and implementing actions. A few NSCTs are assessing the impact of these actions, which actions result in the greatest impact is still unknown. Asking trusts to assess the impact of actions taken, without assessing the impact of a new patient safety programme could be considered hypocritical.

Consensus about what constitutes effective learning with regards to patient safety needs to be defined and agreed upon. Further support for implementation of the LfDs programme should be undertaken. This needs to include a more structured reporting framework. Evidence of effective learning, sharing of learning and engagement with bereaved families should be added to the statutory reporting requirements

The lack of success with the LfDs programme needs to be investigated further.

## Data Availability

All data used in this research is publicly available from NHS Secondary Care Trust websites in their Quality Accounts

## Acknowledgements

We would like to acknowledge the work of the Learning from Deaths: Learning and Action (LfDLaA) Public and Relatives steering group in this research. Patient and public involvement in this research was supported by the NIHR UCL Biomedical Research Centre.

## Supplemental page 1

### Reflexivity detail

The primary reviewer (ZB) is a frontline clinician, who felt embarrassment and disappointment during the collection and analysis phases of this research, by the lack of attention to detail, disregard for undertaking statutory requirements and often indifference demonstrated by NSCTs. It appeared to ZB that some NSCTs see the LfDs programme as another regulatory requirement and not as an opportunity to improve care for future patients. ZB understands the challenges of working in frontline healthcare and appreciates that NSCTs have many competing programmes and issues to manage. ZB shared these reflections and concerns during meetings with the other authors, this process of team-based discussion helped identify assumptions and cross check interpretations.

## Supplemental page 2

### ‘What does ‘learning’ mean?’

Even an understand of what is meant by learning is variable, one NSCT lists ‘learning’ in a seemingly nebulous format, with no further details describing these topics:

‘*We have disseminated learning on a number of thematic lessons using a modality of communication systems:*

- *Sepsis Care Bundles*
- *Fluid management*
- *Appropriate management of pleural effusion’*

A different NSCT does not describe any learning, but instead states: *‘The learning from these is utilised for quality improvement projects’* Learning is sometimes unrelated to patient deaths or even clinical care, as demonstrated by one NSCT LfDs report:

*‘Here are some of the things that we have learnt and implemented:*

- *Quality Academy - Clinical Coding project - Two Junior Doctors (SAMP) to investigate the ‘R’ Code (signs & symptoms) coding issue*.
- *Prioritised Coding – Bereavement notes are prioritised for coding which supports the mortality review process*.*’*

While NSCTs do realise that they are required to ‘learn’ it does not appear that they necessarily understand what effective learning from deaths is or how to describe it in any detail. In addition, ‘Learning’, actions and assessment of impact are frequently merged together, resulting in confusing and amorphous reporting. Another feature of the LfDs reports from some NSCTs is the overwhelmingly ‘positive’ learning, with some NSCTs describing only positive learning despite reporting deaths due to problems in care: ‘*Feedback from bereaved relatives is overwhelmingly positive with 30% of all compliments being sent as excellence reports to ward staff and named individuals*.*’* Of the question of whether learning is actually happening, one NSCT stated: *‘In relation to learning from the structured judgement review processes the following has been highlighted and replicate data from last year’* Implying that the same problems keep reoccurring.

### ‘Undertaking a thematic analysis’

Some NSCTs have undertaken grouping of ‘common themes’ arising and described this as a ‘thematic analysis’, none have given details about the methodology used. The ‘themes’ (domains) arising from these ‘analyses’ are in line with those from this larger review and earlier reports as described in the introduction. One example of this: *‘Incident themes: Communication, Clinical management, Review & escalation, Process & policy, Documentation, Operational, Medication’* However the ‘thematic analysis’ in itself can make ‘learning’, especially shared learning more difficult, due to reduction in detail given in some (but not all) reports.

### ‘Opportunities to triangulate’

Several NSCTs have missed the opportunity to bring together information which could inform learning in their LfDs report. For example reporting no deaths due to problems in care despite being issued PFD reports and/or not describing learning from the PFD reports in their LfDs report. One example from a PFD report describes problems with acute physical assessment and examination in the Mental Health setting. The death of this patient and subsequent learning from the Coroner’s office is not acknowledged in either the expected LfD report.[32] Other NSCTs do acknowledge PFD reports in their LfDs report, but don’t necessarily explain what if anything they have learnt from this.

Some NSCTs do appear to triangulate data, for example learning acquired through the Dr Foster Diagnosis Alerts or associated with HSMR and/or SHMI. One NSCT noted ‘*The themes that emerge from SJRs are representative of the themes and trends that are seen in complaints, incidents and claims’* and another describes ‘*The trust’s Datix incident reporting system is aligned to the Learning from Patient Deaths Policy’*

In NSCTs where Quality Improvement is fully embedded in their standard functioning have been able to successfully tie in LfDs with other work: ‘*Strong inter-relationship of our SI, SJR and improvement programmes, for example our VTE work where SJR and root cause analysis processes have helped to identify key challenges and drive forward improvement’* thus ensuring greater success of the programme and benefits for patients.

### ‘Feeling the pressure’

Systems issues and lack of resources feature surprisingly infrequently in LfDs reports, however in 2019/20 one NSCT explicitly stated that these factors were problematic: ‘*Overload is a significant theme to the cases that have been noted. It is typically present as a contributory factor rather than the only issue. It links to NIV beds, to recognition of pelvic bleeds and to extended stay in A/E. Its also linked to omitted drugs and to sepsis care. When considered across organisations its also linked to complex system failure where beds are too full to receive transfers’* Another NSCT reported issues with bed availability for Mental Health patients: *‘The service transformation teams continue to work in relation to bed capacity and demand which aims to introduce a crisis house and an urgent care centre which should impact upon the prioritisation of beds’*

### ‘Description of the incident/problem’

One of the main issues found from reviewing LfDs reports was the difficulty in understanding what exactly the problem in care was, that may have contributed to the patients death. Very few trusts described individual incidents/cases, but where they did it enables better understanding of what happened, thus enabling shared learning:

- Incident – ‘*death from haemopericardium caused by dissection of the ascending aorta*.
  - Learning/action - *Improve awareness of this rare diagnosis among all ED staff through ongoing teaching sessions and safety briefings*.*’*
- Incident – ‘*There was a missed diagnosis of small bowel obstruction in a patient resulting in aspiration and cardiac arrest, with unsuccessful resuscitation’*
  - Learning – ‘*The missed diagnosis was due to lack of recognition of the significance of symptoms of pain and persistent vomiting in the context of reassuring National Early Warning System (NEWS) scores and apparent initial response to treatment’*
  - Action – *‘As a result of this incident new guidelines for the management of small bowel obstruction have been completed. Processes for recognition and treatment will be embedded into local assessments and practice. Training in the diagnosis and initial treatment of acute surgical conditions and new guidelines’*
- Incident – *‘There have been some cases where we have not detected intra-uterine growth retardation (IUGR), and so have not carried out the correct monitoring and support’*
  - Learning – *‘This issue has arisen due to poor scan quality in some cases, and due to interpretation biases arising from human factors in other cases’*
  - Action – *‘We are carrying out an audit of missed IUGR to inform future practice’*

One NSCT laid out the incident/learning/actions in tabular format, making what happened, what has been learnt and what has occurred as a result of that learning (action) clear.

### ‘The importance of culture’

Very few NSCTs remarked upon the importance of improving culture or providing a ‘Just Culture’:

- *‘By building and nurturing an improved culture, new ways of thinking and working can be introduced, but these new ways will only become embedded within the team if they enable people to work more effectively than before’*

*‘To ensure that the mortality review process leads to meaningful and effective actions that continually improve patient safety and experience operating within a ‘Just Culture’ framework’*

## Supplemental page 3

### Quality Accounts with quotes in paper

- Pennine care NHS Foundation trust Quality Account 2019/20
- Cumbria, Northumberland, Tyne & Wear NHS Foundation Trust Quality Account 2019/20
- North Middlesex University Hospital NHS Trust Quality Account 2018/19
- Nottinghamshire Healthcare NHS Foundation Trust Quality Account 2019/20
- Cambridge University Hospitals NHS Foundation Trust Quality Account 2019/20
- Chelsea and Westminster Hospital NHS Foundation Trust Quality Account 2019/20
- University Hospitals Plymouth NHS Trust Quality Account 2019/20
- Avon and Wiltshire Mental Health Partnership NHS Trust Quality Account 2019/20
- University Hospitals Birmingham NHS Foundation Trust Quality Account 2018/19.
- Gloucestershire Care Services NHS Trust Quality Account 2018/19
- King’s College Hospital NHS Foundation Trust Quality Account 2019/20
- Bradford District Care NHS Foundation Trust Quality Account 2018/19
- Worcestershire Health and Care Quality Account 2017/18
- Harrogate and District NHS Foundation Trust Quality Account 2019/20
- University Hospitals of Derby and Burton NHS Foundation Trust Quality Account 2018/19
- Coventry and Warwickshire Partnership NHS Trust Quality Account 2018/19
- Royal Devon and Exeter NHS Foundation Trust Quality Account 2018/19
- Devon Partnership NHS Trust Quality Account 2017/18
- York Teaching Quality Account 2018/19
- Liverpool Heart and Chest Quality Account 2018/19
- Liverpool Heart and Chest Quality Account 2019/20
- Moorfields Quality Account 2019/20
- Birmingham & Solihull MHT Quality Account 2019/20
- Sussex Partnership Quality Account 2019/20
- Taunton & Somerset Quality Account 2019/20
- Epsom Quality Account 2019/20
- Gateshead Health Quality Account 2019/20
- Northumbria Healthcare Quality Account 2019/20
- Pennine Care Quality Account 2019/20
- Greater Man West MHT Quality Account 2019/20
- Western Sussex Quality Account 2019/20
- Portsmouth Quality Account 2019/20
- ULH Quality Account 2019/20
- Basildon Quality Account 2019/20
- Warrington Quality Account 2019/20
- Buckinghamshire Quality Account 2019/20
- East & North Herts Quality Account 2019/20
- Pennine Care Quality Account 2019/20
- Midlands Partnership Quality Account 2018/19
- Midlands Partnership Quality Account 2019/20
- Wirral Community Quality Account 2019/20
- Western Sussex Quality Account 2019/20
- Bradford Teaching Quality Account 2019/20
- WWL Quality Account 2019/20
- Birmingham and Solihull MHT Quality Account 2019/20
- York Teaching Quality Account 2019/20
- University College London Hospitals NHS Foundation Trust Quality Account 2019/20
- BWCH Quality Account 2019/20
- Hampshire Hospitals Quality Account 2019/20
- St Helens Quality Account 2019/20
- Cornwall Partnership Quality Account 2019/20

## References

1. Brummell Z, Vindrola-Padros C et al. NHS ‘Learning from Deaths’ reports: a qualitative and quantitative document analysis of the first year of a countrywide patient safety programme. BMJ Open 2021;11:e046619. https://doi.org/10.1136/bmjopen-2020-046619

2. National Academies of Sciences, Engineering and Medicine. 2018. Crossing the Global Quality Chasm: Improving Health Care Worldwide. Washington, DC: The National Academies Press. https://doi.org/10.17226/25152

3. Berwick D (2013) A promise to learn – a commitment to act: improving the safety of patients in England. London: Department of Health. Available: https://assets.publishing.service.gov.uk/government/uploads/system/uploads/attachment_data/file/226703/Berwick_Report.pdf

4. Francis R (2013) Report of the Mid Staffordshire NHS Foundation Trust Public Inquiry. London: The Stationery Office. Available: https://assets.publishing.service.gov.uk/government/uploads/system/uploads/attachment_data/file/279124/0947.pdf

5. Kirkup B (2015). The Report of the Morecambe Bay Investigation. London: The Stationary Office. Available: https://assets.publishing.service.gov.uk/government/uploads/system/uploads/attachment_data/file/408480/47487_MBI_Accessible_v0.1.pdf

6. House of Commons Justice Committee Oral evidence: The Coroner Service, HC 282, 2020. Available: https://committees.parliament.uk/oralevidence/1092/html/

7. Titcombe, J. Joshua’s Story: Uncovering the Morecambe Bay NHS scandal. (2015) Anderson Wallace Publishing, UK

8. NQB. Learning from deaths: guidance for NHS trusts on working with bereaved families and carers, 2018. Available: https://www.england.nhs.uk/wp-content/uploads/2018/08/learning-from-deaths-working-with-families-v2.pdf

9. Mazars 2015. Independent review of deaths of people with a Learning Disability or Mental Health problem in contact with Southern Health NHS Foundation Trust April 2011 to March 2015. Available: https://www.england.nhs.uk/south/wp-content/uploads/sites/6/2015/12/mazars-rep.pdf

10. Care Quality Commission 2016. Learning candour accountability: A review of the way NHS trusts review and investigate the deaths of patients in England. Available: https://www.cqc.org.uk/sites/default/files/20161213-learning-candour-accountability-full-report.pdf

11. NQB. National guidance on learning from deaths, 2017. Available: https://www.england.nhs.uk/wp-content/uploads/2017/03/nqbnational-guidance-learning-from-deaths.Pdf

12. NHS Improvement. Implementing the learning from deaths framework: key requirements for trust boards, 2017. Available: https://www.england.nhs.uk/wp-content/uploads/2021/07/170921-Implementing-LfD-information-for-boards.pdf

13. UK Legislation. The National health service (quality accounts) (Amendment) regulations, 2017. Available: https://www.legislation.gov.uk/uksi/2017/744/regulation/2/made

14. NHS. About quality accounts, 2019. Available: NHS. About quality accounts, 2019. Available: https://www.nhs.uk/using-the-nhs/about-the-nhs/quality-accounts/about-quality-accounts/

15. Lalani M, Hogan H. A narrative account of the key drivers in the development of the Learning from Deaths policy. Journal of Health Services Research and Policy 2021. 26(4):263–271. https://doi.org/10.1177/13558196211010850

16. The Coroners (Investigations) Regulations 2013. Available: https://www.legislation.gov.uk/uksi/2013/1629/regulation/29/made

17. Leary A, Bushe D et al. A thematic analysis of the prevention of future deaths reports in healthcare from HM coroners in England and Wales 2016–2019. J Patient Safety & Risk management 2021. 26(1): 14–21

18. National Confidential Enquiry into Patient Outcomes and Deaths (NCEPOD). An acute problem? 2005. Available: https://www.ncepod.org.uk/2005report/NCEPOD_Report_2005.pdf

19. Healthcare Safety Investigation Branch. Health Service Investigation Summary of Themes arising from the Healthcare Safety Investigation Branch Maternity Programme (NLR) 2020. Available: https://hsib-kqcco125-media.s3.amazonaws.com/assets/documents/hsib-national-learning-report-summary-themes-maternity-programme.pdf

20. MBRRACE-UK. Saving Lives, Improving Mothers’ Care: Lessons learned to inform maternity care from the UK and Ireland Confidential Enquiries into Maternal Deaths and Morbidity 2017-19, 2021. Available: https://www.npeu.ox.ac.uk/assets/downloads/mbrrace-uk/reports/maternal-report-2021/MBRRACE-UK_Maternal_Report_2021_-_FINAL_-_WEB_VERSION.pdf

21. Schiff G, Shojanian KG. Looking back on the history of patient safety: an opportunity to reflect and ponder future challenges. BMJ Quality & Safety 2022;31:148–152 https://qualitysafety.bmj.com/content/31/2/148

22. Care Quality Commission. Learning from deaths: A review of the first year of NHS trusts implementing the national guidance 2019. Available: https://www.cqc.org.uk/sites/default/files/20190315-LfD-Driving-Improvement-report-FINAL.pdf

23. Alderson P. Critical Realism for Health and Illness Research: A Practical Introduction. Bristol University Press 2021

24. Bowen GA. Document analysis as a qualitative research method. Qualitative Research Journal 2009;9:27–40

25. https://www.england.nhs.uk/wp-content/uploads/2019/07/learning-from-deaths-guidance-for-ambulance-trusts.pdf

26. Morgan DL. Qualitative content analysis: a guide to paths not taken. Qual. Health Res. 1993; 3: 112–121

27. Braun V, Clarke V. One size fits all? What counts as quality practice in (reflexive) thematic analysis?, Qualitative Research in Psychology 2021, 18(3):328–352, https://doi.org/10.1080/14780887.2020.1769238

28. O’Brien BC, Harris IB, Beckman TJ, et al. Standards for reporting qualitative research: a synthesis of recommendations. Acad Med 2014;89:1245–51

29. Lisa R. Trainor & Andrea Bundon (2021) Developing the craft: reflexive accounts of doing reflexive thematic analysis, Qualitative Research in Sport, Exercise and Health, 13(5):705–726, https://doi.org/10.1080/2159676X.2020.1840423

30. Staniszewska S, Brett J, Simera I, et al. GRIPP2 reporting checklists: tools to improve reporting of patient and public involvement in research. BMJ 2017;358:j3453.

31. Sandelowski M, Leeman J. Writing Usable Qualitative Health Research Findings. Qualitative Health Research 2021;22(10):1404–1413 https://doi.org/10.1177%2F1049732312450368

32. Haigh A. Regulation 28 Report to Prevent Future Deaths. 2019 Available: https://www.judiciary.uk/wp-content/uploads/2019/11/Maureen-Jarvis-2019-0357_Redacted.pdf

33. BBC News. Mothers who helped uncover the biggest NHS maternity scandal. 2022. Available: https://www.bbc.co.uk/news/health-60434299

34. BBC News Essex. Essex mental health trust: Partner ‘begged’ for help before man’s death 2022. Available: https://www.bbc.co.uk/news/uk-england-essex-60402331

35. Waring J, Currie G. The politics of learning: The dilemma for patient safety. A Socio-Cultural perspective on patient safety. CRC Press 2017

36. House of Commons. Health and Social Care Committee. Workforce burnout and resilience in the NHS and social care. Second Report of Session 2021-22. 2021. Available: https://committees.parliament.uk/publications/6158/documents/68766/default/

37. Tabrizi NM, Masri F. Towards safer healthcare: qualitative insights from a process view of organisational learning from failure. BMJ Open 2021; 11:e048036. Available: https://bmjopen.bmj.com/content/bmjopen/11/8/e048036.full.pdf

38. WHO. Global Patient Safety Action Plan 2021. Available: https://www.who.int/publications/i/item/9789240032705

39. HSIB. National learning report. A thematic analysis of HSIB’s first 22 national investigations. September 2021. Available: https://www.hsib.org.uk/

40. Farenden S et al. Impact of implementation of the National Early Warning Score on patients and staff. British Journal Hospital Medicine 2017;78(3). Available: https://www.magonlinelibrary.com/doi/epub/10.12968/hmed.2017.78.3.132

41. Maharaj R et al. Rapid response systems: a systematic review and meta-analysis. Critical Care 2015;19:254. Available: https://link.springer.com/article/10.1186/s13054-015-0973-y

42. Brummell Z et al. Data, clinical coding and clinicians: lost in translation. British Journal of Hospital Medicine 2019;80:7 Available: https://www.magonlinelibrary.com/doi/epub/10.12968/hmed.2019.80.7.364

43. UK Government. Wachter RM. Making IT Work: Harnessing the Power of Health Information Technology to Improve Care in England. 2016. Available: https://assets.publishing.service.gov.uk/government/uploads/system/uploads/attachment_data/file/550866/Wachter_Review_Accessible.pdf

44. O’Hara R, Coster J, Goodacre S. Qualitative exploration of the Medical Examiner role in identifying problems with the quality of patient care. BMJ Open 2021;11:e048007. Available: https://www.ncbi.nlm.nih.gov/pmc/articles/PMC7925852/pdf/bmjopen-2020-048007.pdf

45. Gandhi TK, Kaplan GS et al. Transforming concepts in patient safety: a progress report. BMJ Quality & Safety 2018;27:1019–1026. Available: https://qualitysafety.bmj.com/content/qhc/27/12/1019.full.pdf

46. Lame G, Dixon-Woods M. Using clinical simulation to study how to improve quality and safety in healthcare. BMJ Simulation & Technology Enhanced Learning 2018;6(2):87–94. Available: https://www.ncbi.nlm.nih.gov/pmc/articles/PMC7056349/pdf/bmjstel-2018-000370.pdf

47. Nieva VF, Murphy R et al. From Science to Service: A Framework for the Transfer of Patient Safety Research into Practice. In: Henriksen K et al., editors. Advances in Patient Safety: From Research to Implementation (volume 2: concepts and methodology). Rockville (MD): Agency for Healthcare Research and Quality (US); 2005. Available: https://www.ncbi.nlm.nih.gov/books/NBK20521/#A3538

48. Zipperer L, editor. Patient Safety: Perspectives on Evidence, Information and Knowledge Transfer. Routledge; 2016

49. Karem M, Brault I et al. Comparing interprofessional and interorganisational collaboration in healthcare: A systematic review of the qualitative research. Int J of Nursing Studies 2018;79: 70–83. Available: https://www.sciencedirect.com/science/article/pii/S0020748917302559

50. The King’s Fund. Time to think differently: Competition and choice, 2012. Available: https://www.kingsfund.org.uk/publications/reforming-nhs-within/competition-and-choice

51. Department of Health and Social Care. Health and Care Bill: competition, 2022. Available: https://www.gov.uk/government/publications/health-and-care-bill-factsheets/health-and-care-bill-competition

52. Vindrola-Padros C, Ledger J et al. The Implementation of Improvement Interventions for “Low Performing” and “High Performing” Organisations in Health, Education and Local Government: A Phased Literature Review. International Journal of Health Policy and Management 2020, x(x),1–9. Available: https://www.ijhpm.com/article_3943_bce8f4ecdbb3cc38176dbbf335cea359.pdf

53. Veenstra GL, Ahaus K et al. Rethinking clinical governance: healthcare professional’ views: a Delphi study. BMJ Open 2017;7:e012591. Available: https://bmjopen.bmj.com/content/bmjopen/7/1/e012591.full.pdf

54. Health Services and Delivery Research, No. 4.4. Mannion R, Freeman T, Millar R, et al. Effective board governance of safe care: a (theorectically underpinned) cross-sectioned examination of the breadth and depth of relationships through national quantitative surveys and in-depth qualitative case studies. Southampton (UK): NIHR Journals Library; 2016. Available: https://www.ncbi.nlm.nih.gov/books/NBK338883/

55. Skivington K, Matthews L et al. A new framework for developing and evaluating complex interventions: update of Medical Research Council guidance. BMJ 2021;374:l2061. Available: https://www.bmj.com/content/374/bmj.n2061

56. NHS Resolution. Annual report and accounts 2020/21: A Summary 2021. Available: https://resolution.nhs.uk/wp-content/uploads/2021/12/Annual-report-and-accounts-20-21-summary.pdf

57. Papanicolas I, Mossialos E et al. Performance of UK National Health Service compared with other high income countries: observational study. BMJ 2019;367:I6326. Available: https://www.bmj.com/content/367/bmj.l6326

58. Gardner T, Fraser C. Longer waits, missing patients and catching up. The Health Foundation 2021. Available: https://www.health.org.uk/news-and-comment/charts-and-infographics/how-is-elective-care-coping-with-the-continuing-impact-of-covid-19

59. Health Service Journal. HSJ Patient Safety awards 2017: best organisation, 2017. Available: https://www.hsj.co.uk/patient-safety/hsj-patient-safety-awards-2017-best-organisation/7018911.article

